# Team contact sports in times of the COVID-19 pandemic- a scientific concept for the Austrian football league

**DOI:** 10.1101/2020.11.06.20226977

**Authors:** Antje van der Zee-Neuen, Dagmar Schaffler-Schaden, Jürgen Herfert, James O’Brien, Tim Johansson, Patrick Kutschar, Alexander Seymer, Stephan Ludwig, Thomas Stöggl, David Keeley, Herbert Resch, Jürgen Osterbrink, Maria Flamm

## Abstract

**Background:** Since the beginning of the COVID -19 pandemic, many contact sport teams are facing major challenges to safely continue training and competition.

**Objective:** We present the design and implementation of a structured monitoring concept for the Austrian national football league

**Methods:** 146 professional players from five clubs of the professional Austrian football league were monitored for a period of 12 weeks. Subjective health parameters, PCR- test results and data obtained from a geo-tracking app were collected. Simulations modelling the consequences of a COVID-19 case with increasing reproduction number were computed.

**Results:** No COVID-19 infection occurred during the observation period in the players. Infections in the nearer surroundings lead to increased perceived risk of infection. Geo tracking was particularly hindered due to technical problems and reluctance of users. Simulation models suggested a hypothetical shut-down of all training and competition activities.

**Conclusions:** A structured monitoring concept can help to continue contact sports safely in times of a pandemic. Cooperation of all involved is essential.

**Trial registration:** ID: DRKS00022166 15/6/2020 https://www.who.int/ictrp/search/en/

**Key Points:**

- The results of this study can inform the development of future prevention and monitoring strategies in professional football and beyond, potentially serving as a blueprint for the safe continuation of sports and physical activity, across a broad range of settings, during and following a pandemic such as COVID-19.
- Health parameters should be digitally recorded and closely monitored to enable quick response in case of a COVID-19 infection.

## 1. Introduction

Since the global outbreak of the COVID-19 pandemic, team contact sports are facing major challenges to continue training and competing. Medical recommendations to prevent virus transmission are changing on a day-to-day basis and although physical distancing is currently a widely accepted preventive measure, its application is not feasible in team contact sports [1]. Shared training facilities and close, prolonged contact with opponents and team members can increase transmission risk in these settings.

After a period of home-confined training, professional football teams all around the world have resumed training and competition. Recently, Austria has experienced a renewed increase in cases, but fortunately the intensive care capacities are not exhausted [2]. Following a stabilisation in infection rates, many European countries eased strict measures to prevent COVID-19 transmission in summer 2020, including resuming public activities, reopening borders and increasing the accessibility of public places. Nevertheless, the upcoming winter and increased travel to international games pose new complex challenges for professional sport organisations.

Recent data suggest that the majority of infected people show no symptoms, which hampers identification [3]. As young and fit athletes may well be asymptomatic in the case of an infection, strict management in team contact sports context is warranted. In order to minimise transmission risk and limit potential strain on health services, the resumption of team sports necessitates a clearly defined, well-coordinated prevention strategy. Several concepts for safely resuming national and international competition have been published [4, 5].

On May 12^th^ 2020, the Austrian government granted permission for the resumption of the Austrian national male association football league, on the condition of no spectators and teams adhering to a pre-defined prevention concept. A further condition was scientific monitoring of the resumption process. The authors were involved in designing and implementing this monitoring concept in selected teams. The overarching goal of the prevention concept and scientific evaluation was to allow safe resumption of training and competition through comprehensive health monitoring and tracking of players’ movements both in- and outside the trainings and competition facilities.

The current manuscript reports the design of the monitoring concept, the barriers encountered during the implementation process and also presents simulation models for infections. In addition to describing the monitoring parameters and their variation across the observation period, a statistical estimation of consequences of a rise in COVID-19 basic reproduction number (R(0)) is provided. The R(0) is an indicator of the number of people infected by one person [6]. We simulated the transmission and spread of the virus in a football team when the R(0) rises (e.g. R(0)>1). The design and implementation of the monitoring concept, including the lessons learned can serve as a valuable blueprint for the safe resumption of professional, recreational and educational sports during a pandemic.

## 2. Methods

The monitoring concept was evaluated across a 12-week period from May 15^th^ 2020 to the end of the football season on July 5^th^ 2020. Players from five consenting Austrian professional football clubs (highest two divisions) were included. For reasons of anonymity, clubs were given code-names (A-E).

### 2.1 Instruments and measures

For the monitoring of health and movement outside training/game facilities, the instruments and their content were structured in three categories:

#### Health diaries

- Diaries included several single questions relating to potential COVID-19 symptoms:
- Cough during the last 24 hours (yes/no)
- Breathing difficulties during the last 24 hours (yes/no)
- Loss of sense of smell/taste during the last 24 hours (yes/no)
- Body aches during the last 24 hours (yes/no)
- Proxies for anxiety, namely
  → Sleep quality (10-point scale, 1=bad sleep, problems falling asleep, several night time wake-ups, 10=good sleep, quickly fell asleep slept through the night)
  → Current perceived risk of infection (10-point scale, 1=calm, composed, unconcerned 10=panicked, anxious, severe risk) as a proxy for anxiety related to the pandemic
  → Current perceived recovery state (10-point scale, 1=energetic, cheerful, rested 10=tired, unenthusiastic, exhausted)

#### Objective parameters

- COVID-19 real time polymerase chain reaction (RT-PCR) tests (by means of nasal and/or throat swab) - screening took place once per week and pooled testing was permitted (i.e. in case of positives in one pool, individual tests must be performed). Analyses of samples were performed by laboratories as recommended by the Federal Ministry of the Republic of Austria, Social Affairs, Health, Care and Consumer Protection
- Body temperature (in Celsius) - measured daily by means of a contactless, forehead thermometer
- Oxygen saturation (SpO2) - measured daily by means of a pulse oximeter attached to the left index finger
- Geo-tracking data
- Geo-tracking data allowed monitoring of players’ movement outside the training/competition facilities. This digital parameter was measured using a smartphone application (app), which was specifically designed by the US-based company Electronic Caregiver for the purpose of data collection and installed on players’ primary mobile devices. This app collected motion data triggered by Global Navigation Satellite System (GNSS) data points. The data was stored in flexible schemes of a noSQL database (i.e. data was stored and retrieved without predefined structure), in order to gather the multiple layers of information capture associated with each data point.

Table 1 provides an overview of all parameters, including the mode and frequency of measurement and the individuals collecting the data.

**Table 1.** Overview of included parameters.

| Parameter | Mode of measurements | Frequency of measurement | Rater/Data-collector |
| --- | --- | --- | --- |
| **Health diaries** |  |  |  |
| Sleep quality | Single question, self-perceived rating | Before training or competition | Team physiotherapist |
| Level of recovery | Single question, self-perceived rating | Before training or competition | Team physiotherapist |
| Perceived risk of infection | Single question, self-perceived rating | Before training or competition | Team physiotherapist |
| Loss of sense of smell/taste | Single question, self-perceived rating | Before training or competition | Team physiotherapist |
| Body aches | Single question, self-perceived rating | Before training or competition | Team physiotherapist |
| Cough | Single question, self-perceived rating | Before training or competition | Team physiotherapist |
| Difficulties in breathing | Single question, self-perceived rating | Before training or competition | Team physiotherapist |
| **Objective parameters** |  |  |  |
| COVID-19 RT-PCR | Throat or nasal swab | Weekly | Team physician |
| Body temperature | Contactless, forehead | Before training or competition | Team physiotherapist |
| Oxygen saturation (SpO<sub>2</sub>) | Pulse oximeter (finger) | Before training or competition | Team physiotherapist |
| **Geo tracking** |  |  |  |
| Geo-tracking data | Smartphone | Continuous | Mobile Application |

### 2.2 Data collection procedures and data quality assurance

During the observation period, standardized data collection sheets were distributed to participating clubs on a weekly basis. Data were collected by team physicians (i.e. COVID-19 RT-PCR tests) and physiotherapists, henceforth referred to as club-internal raters.

A contact person from the Paracelsus Medical University Salzburg (PMU) sent weekly reminders and updates to participating clubs. Three external, student raters, were employed and trained by the researchers to assist the club-internal raters with the collection of health data and the installation of the smartphone app for geo-tracking. The researchers cleaned the data and established a database in preparation, for statistical analysis using R version 3.5.2, R Core Team, 2018.

### 2.3 Statistical analyses and presentation of results

#### Health diary data and objective parameters

For the purpose of the current manuscript, descriptive statistics were employed according to parameters metric properties: Mean and standard deviation (SD) or, where appropriate, median and interquartile range (IQR) for continuous variables and the total number (n) and percentage (%) for categorical variables.

In club A, a management employee, who had no direct contact with the players, tested positive to COVID-19 during the study period. To compare the subjective perceived risk of infection in players from club A before and after this confirmed COVID-19 case, a Wilcoxon-signed-rank test with continuity correction was performed. The test compared the mean score of infection risk per player one week prior to the COVID-19 case to the mean score of infection risk one week after the case.

#### Geo-tracking

To determine whether the range of movement of all included players varied substantially from their expected movements (i.e. in the proximity of their home and training and competition facilities) heat maps were generated from the geo-tracking data

#### Simulation of COVID-19 case and potential consequences according to reproduction number R(0)

The impact of a hypothetical COVID-19 case on the continuation of training and competition in a participating club were simulated for randomly picked R(0)s. For this purpose, 30 players (as a nearly closed cohort) were considered and an observation period of 90 days was assumed. The simulated COVID-19 case assumed the following: (1) the development of symptoms takes (approximately) 5 days, (2) players are tested every 5 days, (3) positively tested players are sent into a quarantine of 14 days, (4) players who had a lot of close contact may be infected or not (i.e. the number of new infected players depends on the R(0), (5) newly infected players may be identified at the subsequent RT-PCR-test, (6) after players return from their quarantine, they do not get infected with COVID-19 again.

For example, we considered simulations for R(0)=1.4 and R(0)=2.15. First, a vector of different R(0) was simulated so that the average R(0) takes the value 2.15 or 1.4, respectively. To account for variation in the number of persons infected by one infected individual (i.e. none, one, > one), a R(0) was drawn randomly from the vector of R(0)s for every player. In order to consider different severities in the course of the spread of the virus, simulations were repeated 100 times for R(0)=1.4 and for R(0)=2.15, respectively.

#### Encountered implementation barriers

Barriers to implementing the monitoring concept, as encountered by the researchers during the study period, were documented and summarized descriptively.

## 3. Results

The full sample consisted of 146 players from five clubs (club A, n=27; club B, n=30; club C, n=28; club D, n=29, club E, n=32).

### Health diary data and objective parameters

Only 4 (14.29%) club C players and 1 (3.45%) club D player reported a sore throat, 2 (7.14%) club C players reported coughing and 1 (3.57%) club C player reported body aches. Health parameters were comparable across clubs (Table 2). The reported and measured health parameters suggest that players from all clubs were in good health during the observation period. None of the players tested positive for COVID-19. However, some variation was found in the parameters sleep quality, level of recovery and perceived risk of infection (Figure 1). In club A, the players’ perceived risk of infection in the week after a manager tested positive for COVID-19 was significantly higher than the perceived risk in the week prior to the occurrence (V=3.5; p<0.05).

**Table 2.** Descriptive statistics of collected health parameters according to club over the entire observation period

|  | Football Clubs |  |  |  |  |
| --- | --- | --- | --- | --- | --- |
|  | Club A<br>n=1091* | Club B<br>n=1277* | Club C<br>n=1173* | Club E<br>n=675* | Club F<br>n=912* |
| **Body Temperature [°C]** |  |  |  |  |  |
| mean (SD) | 36.33 (0.26) | 36.52 (0.21) | 35.86 (0.33) | 36.39 (0.35) | 36.30 (0.35) |
| **Oxygen Saturation [%]** | n(valid)=365 | n(valid)=1009 | n(valid)=227 | n(valid)=297 | n(valid)=351 |
| mean (SD) | 98.45 (0.67) | 98.32 (0.76) | 98.21 (0.69) | 97.77 (1.05) | 97.86 (1.02) |
| **Sleep Quality (1 – 10)** |  |  |  |  |  |
| Median (IQR) | 2 (2-3) | 1 (1-1) | 2 (2-3) | 1 (1-1) | 2 (2-3) |
| **Recovery (1 – 10)** |  |  |  |  |  |
| median (IQR) | 2 (2-3) | 1 (1-1) | 2 (2-4) | 1 (1-1) | 2 (2-3) |
| **Risk of Infection (1 – 10)** |  |  |  |  |  |
| median (IQR) | 1 (1-2) | 1 (1-1) | 1 (1-1) | 1 (1-1) | 1 (1-1) |
| **Sore throat, yes, n\*\* (%)** | 0 (0) | 0 (0) | 15 (1.28) | 1 (0.15) | 0 (0) |
| **Cough, yes, n\*\* (%)** | 0 (0) | 0 (0) | 10 (0.85) | 0 (0) | 0 (0) |
| **Body aches, yes, n\*\* (%)** | 0 (0) | 0 (0) | 1 (0.09) | 0 (0) | 0 (0) |
| **Breathing difficult, yes, n\*\* (%)** | 0 (0) | 0 (0) | 1 (0.09) | 0 (0) | 0 (0) |
| **Loss of taste, yes, n\*\* (%)** | 0 (0) | 0 (0) | 0 (0) | 0 (0) | 0 (0) |
| * denotes the number of data points during the observation period by club<br>**denotes the number of times a specific symptom was reported (during the whole observation period).<br>IQR = Interquartile range (1st quartile & 3rd quartile), SD = standard deviation |  |  |  |  |  |

**Figure 1.**
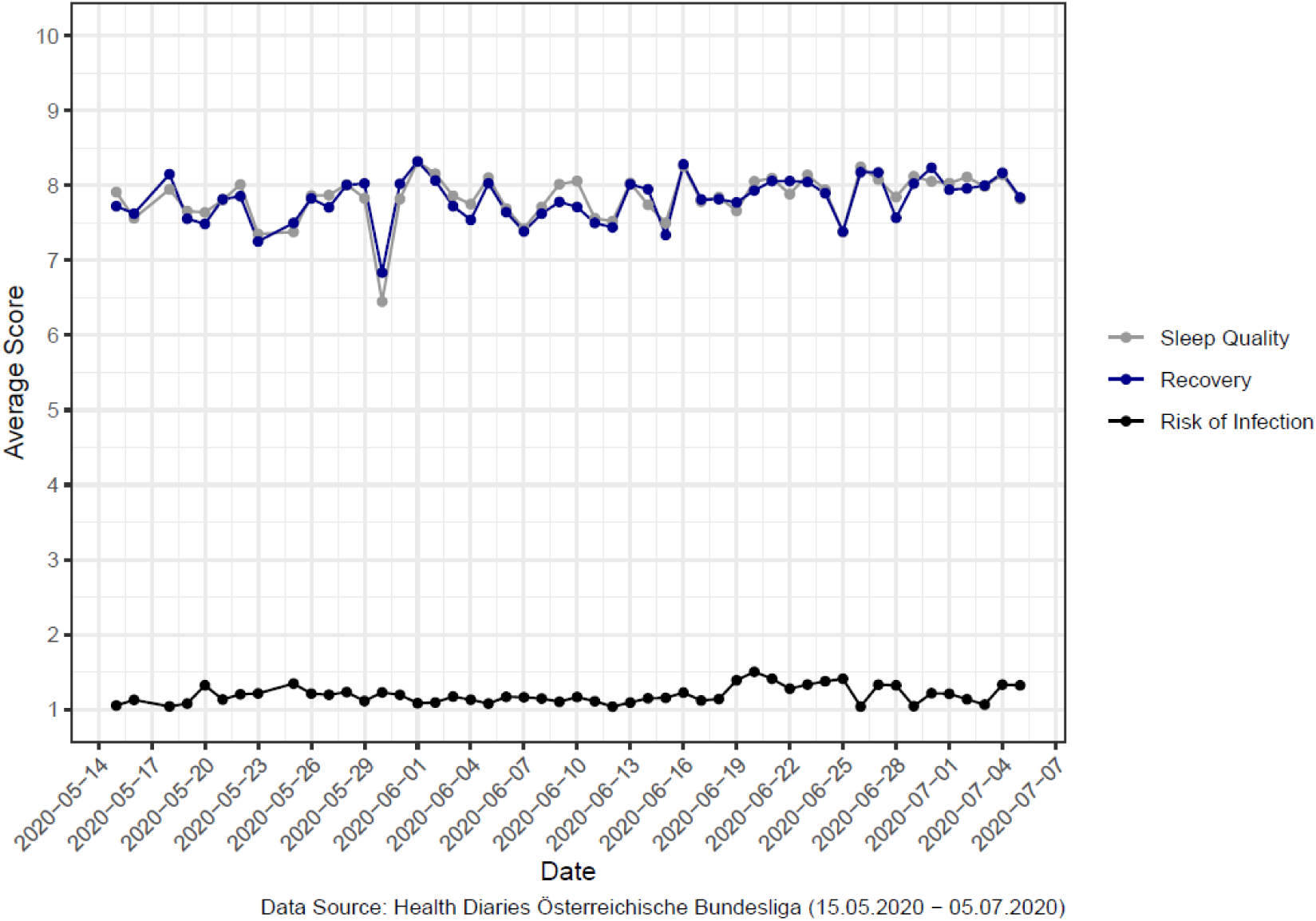
Mean score of perceived sleep quality, level of recovery and risk of infection over time across all participating players (scales 1-10; sleep quality: 1-good sleep – 10-bad sleep; recovery: 1-energetic – 10-tired; risk of infection: 1-calm – 10-anxious)

### Geo-tracking

The collected geo-tracking data suggest that participants’ geographical movement was largely restricted to areas in the proximity of their training facilities and football stadiums they travelled to for the purpose of external matches. (Figure 2)

**Figure 2.**
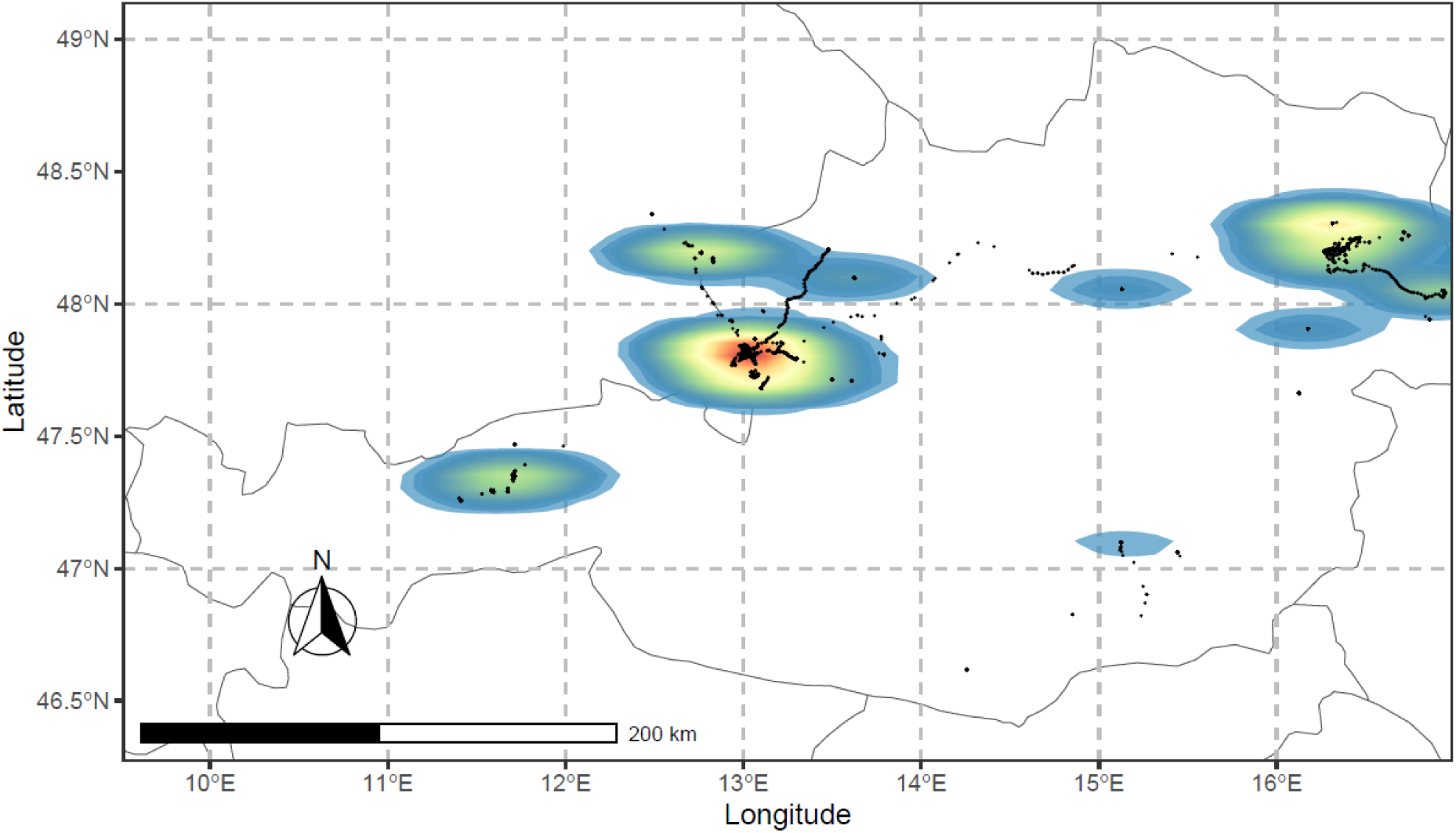
Heat map of collected geo tracking data of participants of five clubs of the Austrian Bundesliga (time period: June 17^th^ 2020 – July 30^th^ 2020). The color gradient can be interpreted as follows: areas with high movement frequencies are displayed with red, areas with less movement frequencies are colored with blue.

### Simulation of COVID-19 case and potential consequences according to reproduction number R(0)

Figure 3 illustrates the proportion of people infected with COVID-19 over a period of 90 days based on the assumptions listed in the method section. As can be seen, the proportion largely depends on the R(0) and differs substantially according to the actual number of infected players. In case of a higher R(0), multiple infections among players could eventually lead to a complete cessation of all training and competitive activities. For instance, in case of R(0)=2.15, after approximately 25 days, about 50% of the player are in quarantine. After 90 days, all players have been infected and have returned from quarantine.

**Figure 3.**
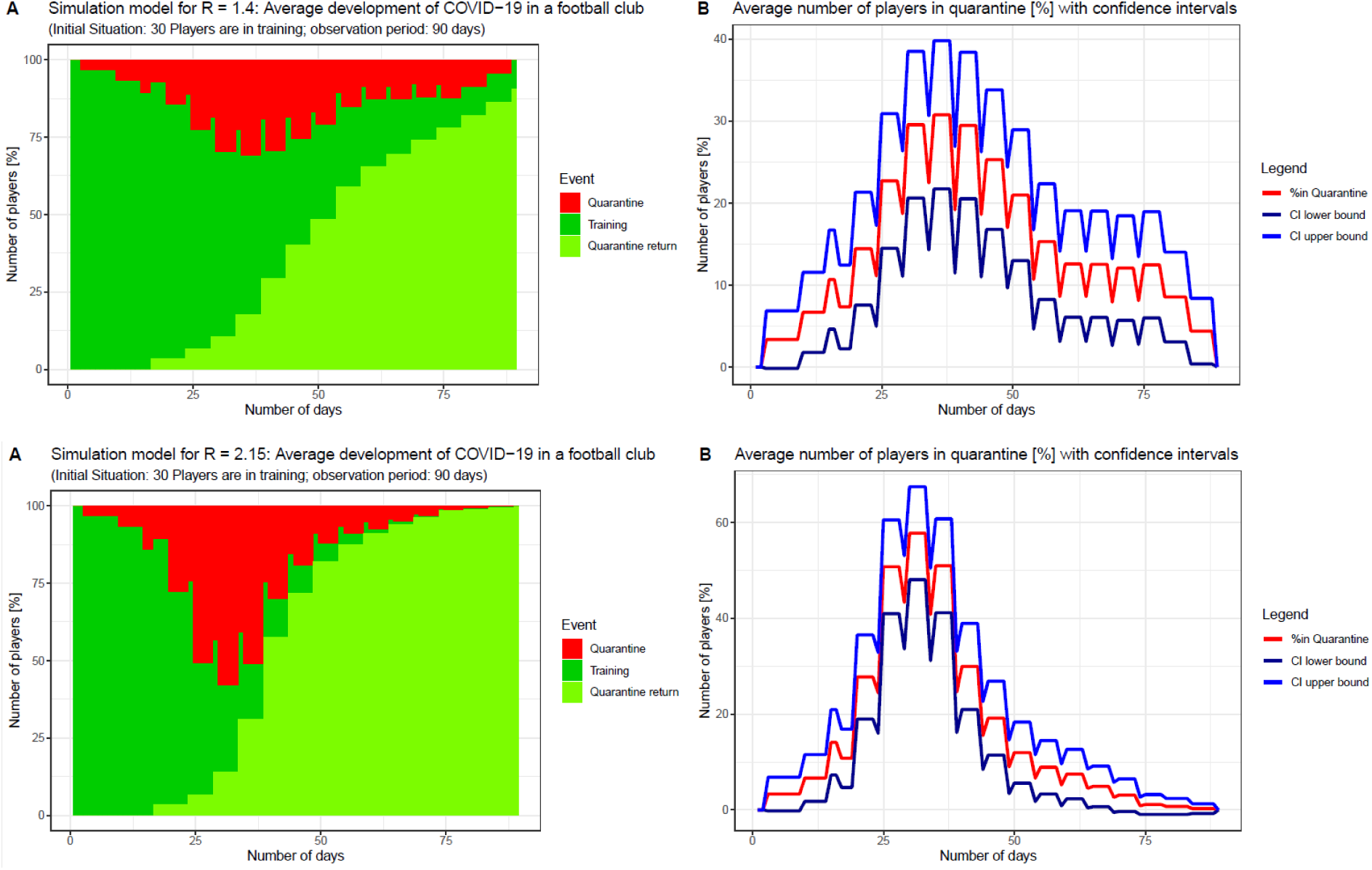
Simulation of hypothetical COVID-19 case among 30 players when R(0)=1.4 or R(0)=2.15

### Encountered implementation barriers

Implementation barriers relating to health parameters were scarce. Data collection sheets were returned weekly to the researcher with the exception of one club, which failed to send information for a period of 2 weeks. Of the 146 players, 5305 data points were generated digitally and 329 data points were generated manually by the club-internal raters. Manually generated data points showed some weaknesses in terms of structuring of collected data. There were 5128 data points with complete information in all parameters (excluding oxygen saturation as obtaining uniform and valid devices was burdensome) and 506 data points with incomplete information. These data points were excluded from statistical analysis. Figure 4 summarizes the completeness of collected data in terms of generated data points.

**Figure 4.**
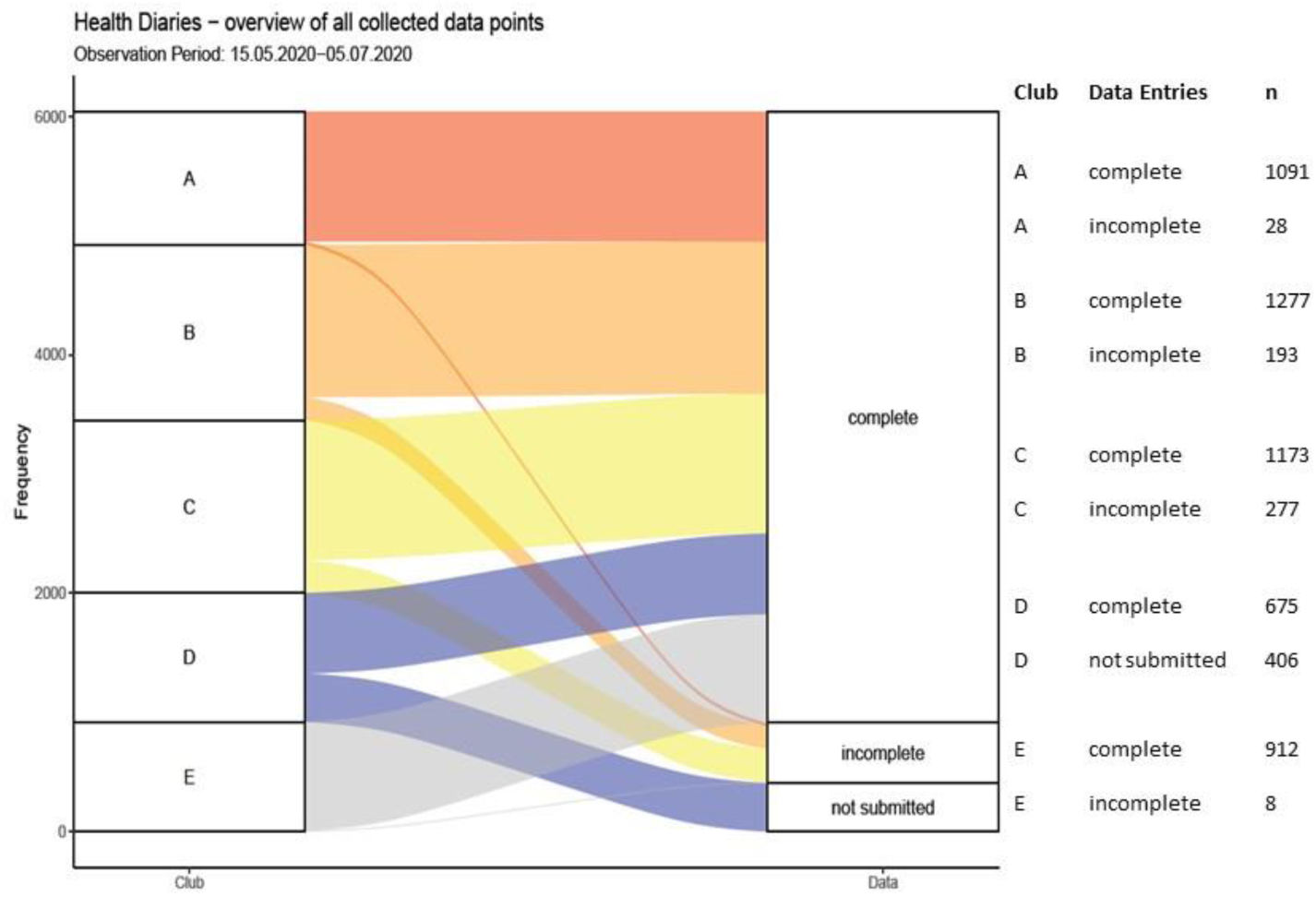
Completeness of health diaries from participating clubs during observation time

The implementation of the smartphone app for the collection of geo-tracking data was more challenging. Internal and external raters reported a reluctance of players to activate geo-tracking and a fear of their movements being monitored by their club. Additionally, technical challenges included unintended shutting down of the app through swiping, installation difficulties on certain smartphones and low density of collected data points occurred. The Flow-chart in Figure 5 depicts the technically-related implementation barriers and their impact on the final sample with geo-tracking data available for analysis.

**Figure 5.**
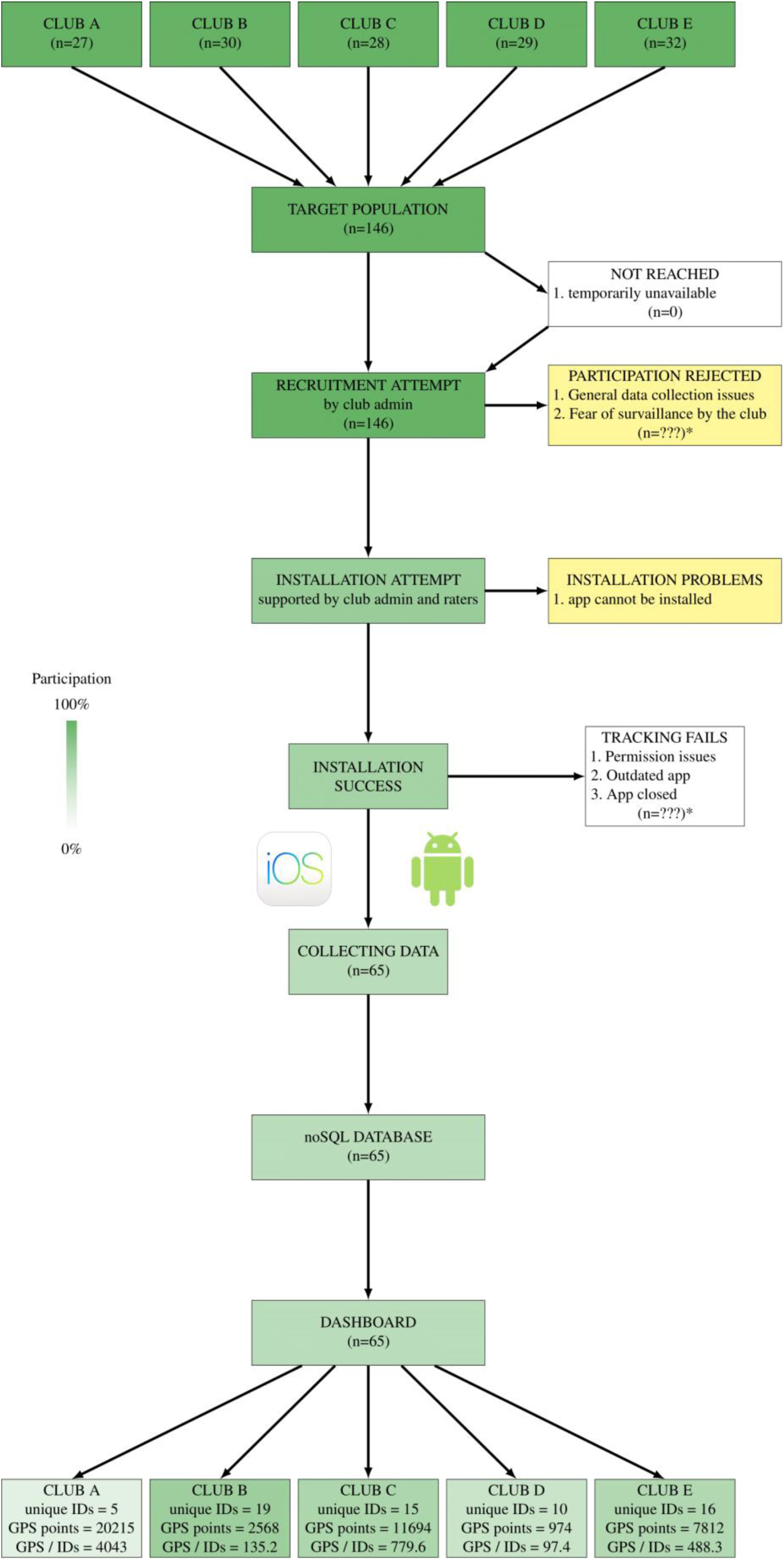
Flowchart of data collection through geo-tracking app and encountered barriers *drop-out not assignable to specific reason, therefore n is unknown

## 4. Discussion

To our knowledge, this is one of the first papers to evaluate the process of safely resuming professional team contact sports during a pandemic. The results of our study suggest that even in times of a pandemic, the continuation of professional football is possible, on the condition of a rigorous surveillance concept. The ongoing pandemic is a challenge for sport clubs all over the world, who are under pressure to continue operating, while ensuring the highest possible level of safety. The quarantining of players can prevent clubs from putting their most important players on the pitch for crucial matches [7].

Physical distancing and contact tracing are currently deemed the most effective strategies to reduce the risk of infection [8]. Regular PCR testing is also crucial for identifying infected persons early, although potential false negatives and the incubation period must be considered [9]. Even with meticulous adherence to all precautionary measures, transmission cannot be completely ruled out. The concept presented in the current study, along with the identification of implementation barriers, can inform future efforts to safely resume team contact sports.

During the observation period, none of the participants tested positive for COVID-19 and variations in health parameters were scarce; only sleep quality and level of recovery displayed slight variation. In line with previous studies reporting impaired sleep quality prior to competitions, these variations are likely attributable to tension and fatigue before and after games [10, 11]. The weekly subjectively perceived risk of infection, which was included as a proxy for anxiety, significantly deviated from the weekly average when a COVID-19 case occurred in a manager in one of the clubs. This deviation was evident despite the fact that players were not in direct contact with the infected individual. This finding is particularly relevant when considering associations between anxiety and performance. Nieuwenhuys et al. [12] suggested that anxiety can affect performance on various levels of operational control and potentially influence perceptual-motor behavior (including situational awareness and decision making). Accordingly, it seems plausible that player performance during competitions might be affected by the subjectively perceived risk of infection. This also highlights the importance of mental support strategies, including regular player education and reassurance, in addition to physical monitoring. Furthermore, adequate interdisciplinary communication (i.e. between management, trainers, physicians/physiotherapists and players) is paramount.

The absence of a COVID-19 case during the observation period of our study could be partially attributable to its low incidence and basic reproduction number in Austria during the observation period [2]. However, players may also travel to other, more affected countries and the situation in Austria is dynamic and evolving. Therefore, we simulated hypothetical scenarios in which the basic reproduction number of the virus rises. These simulations suggest that a worst-case scenario would result in discontinuation of training and competition. While these simulations are based on a number of assumptions, they highlight the necessity of comprehensive and ongoing player monitoring during the pandemic. Even in the case of asymptomatic infections, early detection is crucial to reduce the risk of further transmission and possible infection of vulnerable persons (e.g. relatives). Additionally, the number of players in professional squads is limited, meaning that a high R would end the league prematurely.

Continuous player monitoring is strenuous for data collectors and often an additional burden on top of daily duties. Efficient and user-friendly data collection is an important consideration; we provided digital options for data collection (i.e. a digital dashboard or excel sheets), training and on-site support in the early phase of the study. Despite this, some data entry errors were evident which naturally rendered data analyses more difficult. Accordingly, future monitoring concepts should focus on the training of data collectors to ensure consistent data collection.

The collection of geo-tracking data poses further challenges. Several countries have implemented digital contact tracing apps during the pandemic, mostly on a voluntary basis [13]. Geo-tracking mobile phone apps can potentially aid contact tracing, on the assumption of participants’ consent and compliance with applicable data protection regulations. These technical solutions are designed to enable faster identification and isolation of infected persons, but users’ concerns about privacy and governmental handling of personal data may hinder their application. It has been reported that the acceptance of app-based contact tracing is increased in persons who have comorbidities and always carry their phone [14]. Usually, young people like professional football players have greater technically affinity and should not encounter any difficulties in the installation of smartphone applications. However, in our study we encountered several barriers that hindered the use of the application. Firstly, there were several technical problems, secondly, the data collection appeared to be hindered by reluctance of players regarding geo-tracking, despite their informed consent to participate in the study. The reasons for this could not be established but may be attributable to data privacy concerns. Due to the strict data protection regulations in place and the anonymization of the data throughout the entire study, it was almost impossible to allocate precise numbers on each obstacle during the data collection process. It should be noted that participants of the study were reassured by our study assistants, their trainers, physicians and physiotherapists, that their data would be treated confidentially. The continuation of geo-tracking depends on the willingness of participants to collaborate. The inclusion of participants in the initial phase of monitoring prior to installation of the app may serve as a potential solution. In the context of large sporting events, not only the protection of players, but also the spectators needs to be considered and the use of apps is a potential solution [15]. Moreover, it should be emphasized that strict regulations only make sense if everyone adheres to them. This requires high motivation and cooperation of the whole team as they are also serving as role models for spectators.

Football is a popular sport in Europe, but the results of the study also hold relevance for to other team contact sports (e.g. rugby, basketball or American football) and settings (e.g. recreational and educational). Health parameters should be digitally recorded and closely monitored to enable quick responses. Furthermore, mental health aspects should be included as part of a comprehensive monitoring concept. Mobile phone contact tracing apps can potentially support the identification and containment of infections. The results of this study can inform the development of future prevention and monitoring strategies in professional football and beyond, potentially serving as a blueprint for the safe continuation of sports and physical activity, across a broad range of settings, during and following a pandemic such as COVID-19.

## Data Availability

Original data are stored on the server of the Paracelsus Medical University of Salzburg and are available upon request.

## 5. Acknowledgements

The authors would like to acknowledge the tremendous effort of Melina Mokry who was of great assistance in the organization of the study, the collection of data and the communication with all stakeholders. Moreover, we would like to thank our external raters, Anna Schmuttermair, Anna-Maria Wörndle and Markus Huthöfer, for the on-site assistance in participating clubs, thereby enabling smooth collection of data. We also would like to express our appreciation for the great work of Michael Lyon and Isiah Fielder for their continuous efforts and patience when developing the smartphone app according to our needs and wishes.

## Declarations/ Compliance with Ethical Standards

### Funding

The Centre for Public Health and Health Services Research of the Paracelsus Medical University received funding from the Red Bull Athlete Performance Centre for the scientific accompaniment of the monitoring concept presented in this manuscript.

### Conflict of interest

None declared

### Authors contributions

All authors contributed to the study conception and design. Material preparation, data collection and analysis were performed by Alexander Seymer, Antje van der Zee-Neuen and Dagmar Schaffler-Schaden. The first draft of the manuscript was written by Antje van der Zee-Neuen & Dagmar Schaffler-Schaden and all authors commented on previous versions of the manuscript. All authors read and approved the final manuscript.

### Ethics

The need for ethical approval was waived (statement of the ethics board of Salzburg county, ID 415-EP/73/820-2020), informed consent was provided by all participants

## Notes

### Competing Interest Statement

The authors have declared no competing interest.

### Clinical Trial

DRKS00022166 15/6/2020 https://www.who.int/ictrp/search/en/

